# Gut-bladder axis syndrome associated with recurrent UTIs in humans

**DOI:** 10.1101/2021.11.15.21266268

**Authors:** Colin J. Worby, Henry L. Schreiber, Timothy J. Straub, Lucas R. van Dijk, Ryan A. Bronson, Benjamin Olson, Jerome S. Pinkner, Chloe L. P. Obernuefemann, Vanessa L. Muñoz, Alexandra E. Paharik, Bruce J. Walker, Christopher A. Desjardins, Wen-Chi Chou, Karla Bergeron, Sinéad B. Chapman, Aleksandra Klim, Abigail L. Manson, Thomas J. Hannan, Thomas M. Hooton, Andrew L. Kau, H. Henry Lai, Karen W. Dodson, Scott J. Hultgren, Ashlee M. Earl

## Abstract

Recurrent urinary tract infections (rUTIs) are a major health burden worldwide, with history of infection being a significant risk factor. While the gut is a known reservoir for uropathogenic bacteria, the role of the microbiota in rUTI remains unclear. We conducted a year-long study of women with and without history of rUTIs, from whom we collected urine, blood and monthly fecal samples for multi-omic interrogation. The rUTI gut microbiome was significantly depleted in microbial richness and butyrate-producing bacteria compared to controls, reminiscent of other inflammatory conditions, though *Escherichia coli* gut and bladder dynamics were comparable between cohorts. Blood samples revealed signals of differential systemic immunity, leading us to hypothesize that rUTI susceptibility is in part mediated through a syndrome involving the gut-bladder axis, comprising gut dysbiosis and differential immune response to bacterial bladder colonization, manifesting in symptoms. This work highlights the potential for microbiome therapeutics to prevent and treat rUTIs.

## Introduction

Urinary tract infections (UTIs) are one of the most common bacterial infections worldwide and are a significant cause of morbidity in otherwise healthy females, with uropathogenic *Escherichia coli* (UPEC) being the primary causative agent ^1^. One of the strongest risk factors for UTI is a history of prior UTIs ^2^, but the biological basis for this phenomenon, and the risk factors for long-term recurrence remain unclear in otherwise healthy women. 20-30% of women diagnosed with a UTI will experience a recurrent UTI (rUTI) in the following months, with some suffering six or more per year, but the reason for this is mostly unknown. Over one million women in the United States are referred to urologists each year because of rUTIs, and the rapid spread of antibiotic resistance in uropathogens is making treatment even more challenging. The gut is a reservoir for UPEC and UTIs most commonly arise via the ascension of UPEC from the gut to the urinary tract ^3–5^. Recent studies have explored the ‘gut microbiota-UTI axis’, showing that uropathogen abundance in the gut is a risk factor for UTI in kidney transplant patients ^6^, and that a ‘bloom’ in uropathogen gut abundance may precede infection ^7^. Other studies have demonstrated differences in gut microbiome composition associated with children suffering UTIs ^8^, and with kidney transplant patients developing bacteriuria ^9^, compared to healthy controls. Furthermore, fecal microbiota transplants to treat *Clostridium difficile* infections may have the collateral effect of reducing the frequency of rUTI ^10,11^, suggesting that perturbation of the gut microbiota can modulate rUTI susceptibility.

It is increasingly accepted that the gut microbiota can play a role in conditions affecting distal organs – for instance, the gut-brain axis and the gut-lung axis are the subject of much ongoing research ^12–15^. However, the gut-bladder axis – the spectrum of direct and indirect interactions between gut flora and the bladder immune and/or infection status – remains uncharacterized, and the role of the gut microbiota in rUTI susceptibility is not well understood. In particular, no study has yet been able to ascertain whether: i) gut dysbiosis is associated with rUTI susceptibility; ii) rUTI women have unique uropathogen dynamics within and between the gut and the bladder; or iii) microbiome-mediated immunological differences may be linked to rUTI susceptibility, as seen in a range of other diseases ^16^.

Here, we present results from the UTI microbiome (UMB) project, a year-long clinical study of women with a history of rUTI and a matched cohort of healthy women. Our unique longitudinal study design allowed us to explore the importance and interdependence of the gut microbiota and *E. coli* strain dynamics in rUTI, susceptibility to infection, and host immune responses that may impact these dynamics. Using metagenomic techniques, including newly developed algorithms to track low abundance *E. coli* at the strain-level, we determined that: **i)** compared to healthy controls, women with a history of rUTI had a distinct, less diverse gut microbiota, depleted in butyrate producers and exhibiting characteristics of low-level inflammation; **ii**) differential immunological biomarkers suggest rUTI women may have a distinct immune state, **iii)** *E. coli* strains were transmitted from the gut to the bladder in both cohorts, though no UTI symptoms occurred in healthy controls, and **iv)** UTI-causing *E. coli* strains often persistently colonized the gut, and were not permanently cleared by repeated antibiotic exposure. Thus, susceptibility to rUTI is in part mediated through a syndrome involving the gut-bladder axis, comprising a dysbiotic gut microbiome with reduced butyrate production capacity and apparent alterations of systemic immunity. Our work shows that UPEC strains persist in the gut despite antibiotic treatment, which itself may exacerbate gut dysbiosis. Therapies targeting UPEC in the gut and/or the microbiome, *e*.*g*., therapeutics to attenuate gut dysbiosis, could be promising alternatives to antibiotics.

## Results & Discussion

### Frequent antibiotic exposure and *E. coli* infections in rUTI cohort

Women with a history of rUTI were recruited to the UMB study, along with an age- and community-matched control cohort comprising healthy women (*Methods*). A total of 16 control and 15 rUTI women participated in the year-long study, providing monthly home-collected stool samples, as well as blood, urine and rectal swabs at enrollment and subsequent clinic visits for UTI treatment (Figure 1a). Participants completed monthly questionnaires on diet, symptoms, and behavior (Supp Data). There was a greater proportion of white women in the rUTI cohort, and self-reported antibiotic use was higher in this group in line with UTI treatment; otherwise, few dietary or behavioral differences were apparent (Table S1).

**Figure 1.**
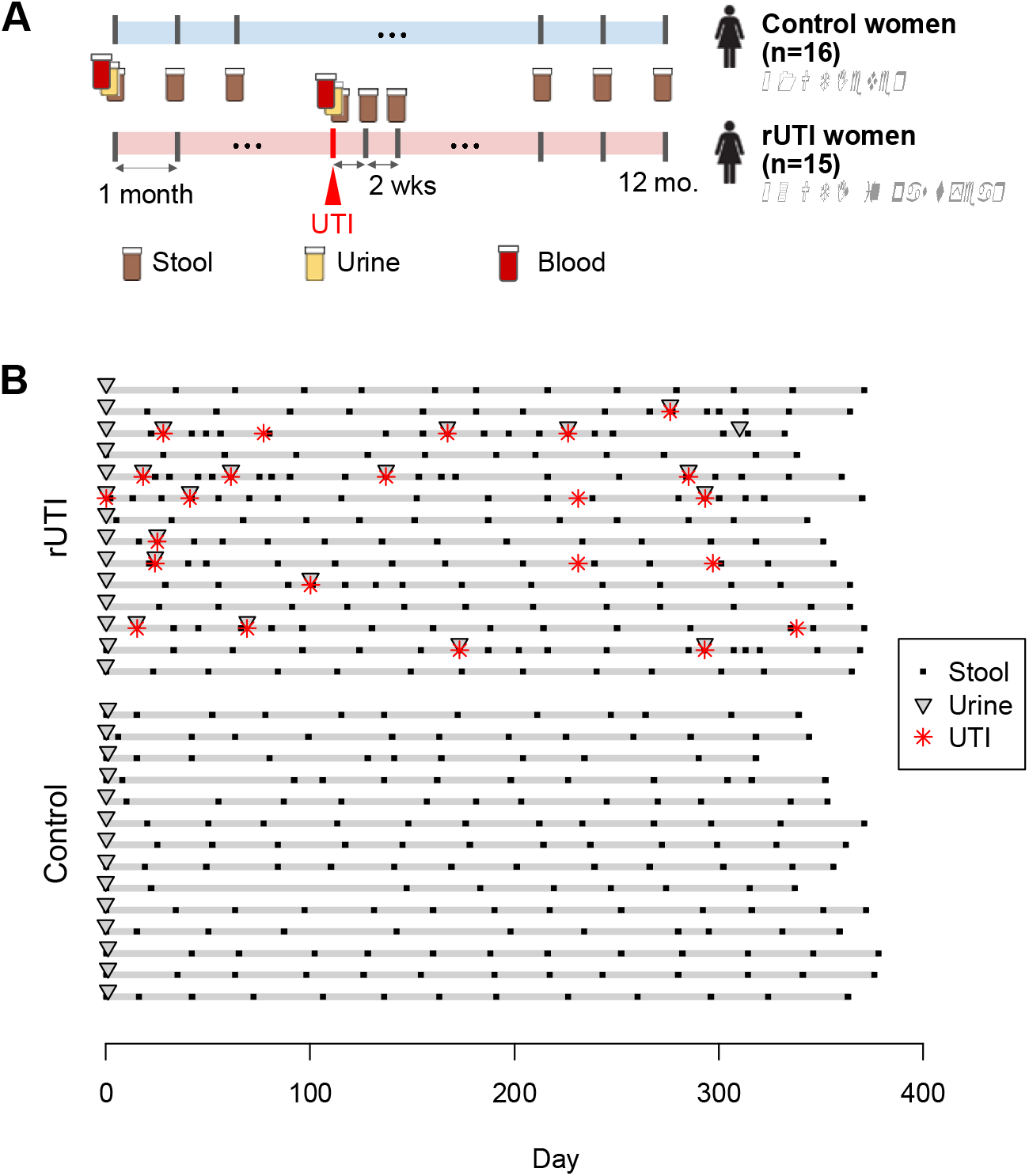
Study design and sample collection for the UMB study. (a) Stool samples were collected monthly from rUTI and control patients. Stool, urine and blood plasma samples were collected upon enrollment and subsequent UTI clinic visits. Biweekly stool samples were requested following UTI diagnoses. (b) Stool and urine samples collected from all rUTI and control participants who completed the study protocol. Red symbols denote diagnosed and inferred UTI events.

A total of 24 UTIs occurred during the study, all occurring in rUTI women, who each experienced between 0-4 UTIs (Figure 1b). Nineteen UTIs were diagnosed by clinicians at the study clinic, and five were inferred through self-reported UTI symptoms (painful urination, increased urgency/frequency of urination, cloudy urine) together with subsequent antibiotic use as reported in the questionnaire administered during monthly sample collection. UTIs were typically treated with ciprofloxacin or nitrofurantoin. No significant temporal risk factors for UTI were identified amongst dietary or behavioral variables. Sexual intercourse is a well-known risk factor for UTI ^2,17^, and all 19 clinically diagnosed UTIs occurred following at least one reported sexual encounter in the previous two weeks (Figure S1).

Urine samples collected at the time of clinical UTI diagnoses were plated on MacConkey agar; bacterial growth was detected (> 0 CFU/ml) from the majority (15/19; 79%, Table S2). To determine the cause of infection, we sequenced 13 urine cultures as well as uncultured urine from all UTI diagnoses, defaulting to results from cultures, when available. *E. coli* dominated 12/13 (92%) sequenced outgrowths; the remaining sample was dominated by *Klebsiella pneumoniae*. Sequencing uncultured urine from the remaining UTI samples identified uropathogens in a further four samples, including *E. coli* (2), *Enterococcus faecalis* and *Staphylococcus saprophyticus*, while two yielded no bacterial sequence (Table S2). Based on sequencing, we defined 14 *E. coli* UTIs, comprising 82% of infections for which a bacterial cause could be inferred, broadly reflecting previous estimates of the proportion of all UTIs caused by *E. coli* ^1^.

### rUTI gut is depleted in microbial richness and butyrate-producing taxa reminiscent of antibiotic exposure

It is increasingly recognized that the gut microbiota plays a role in a range of autoimmune and inflammatory diseases ^18^, as well as susceptibility to infection ^16^, and can alter inflammation in distal organs ^19^. While previous studies have highlighted differential abundances of non-uropathogenic gut taxa as risk factors for bacteriuria in kidney transplant patients (reduced *Faecalibacterium* and *Romboutsia* ^9^) and UTIs in children (reduced *Peptostreptococcaceae* ^8^), it is unclear if these are also risk factors for recurrence in otherwise healthy adult women. To explore this question, we sequenced and analyzed the metagenomes of 367 longitudinal stool samples from both rUTI (n=197) and control (n=170) women (Figure 1b; *Methods*).

After adjusting for potential confounding factors (race and recent antibiotic use), we found broad differences in the composition of the gut microbiota between cohorts (Figure 2a-c). Gut microbial richness was significantly lower, on average, in rUTI women compared to controls (p=0.05, Figure 2c). At the phylum level, we saw elevated levels of *Bacteroidetes* (false discovery rate [FDR]=0.003) and a lower relative abundance of *Firmicutes* (FDR=0.02)) in rUTI women. We identified 22 differentially abundant taxa (FDR<0.25) at lower taxonomic levels, 16 of which were depleted in rUTI women (Table S3; Figure 2b), including *Faecalibacterium* as previously reported ^9^.

**Figure 2.**
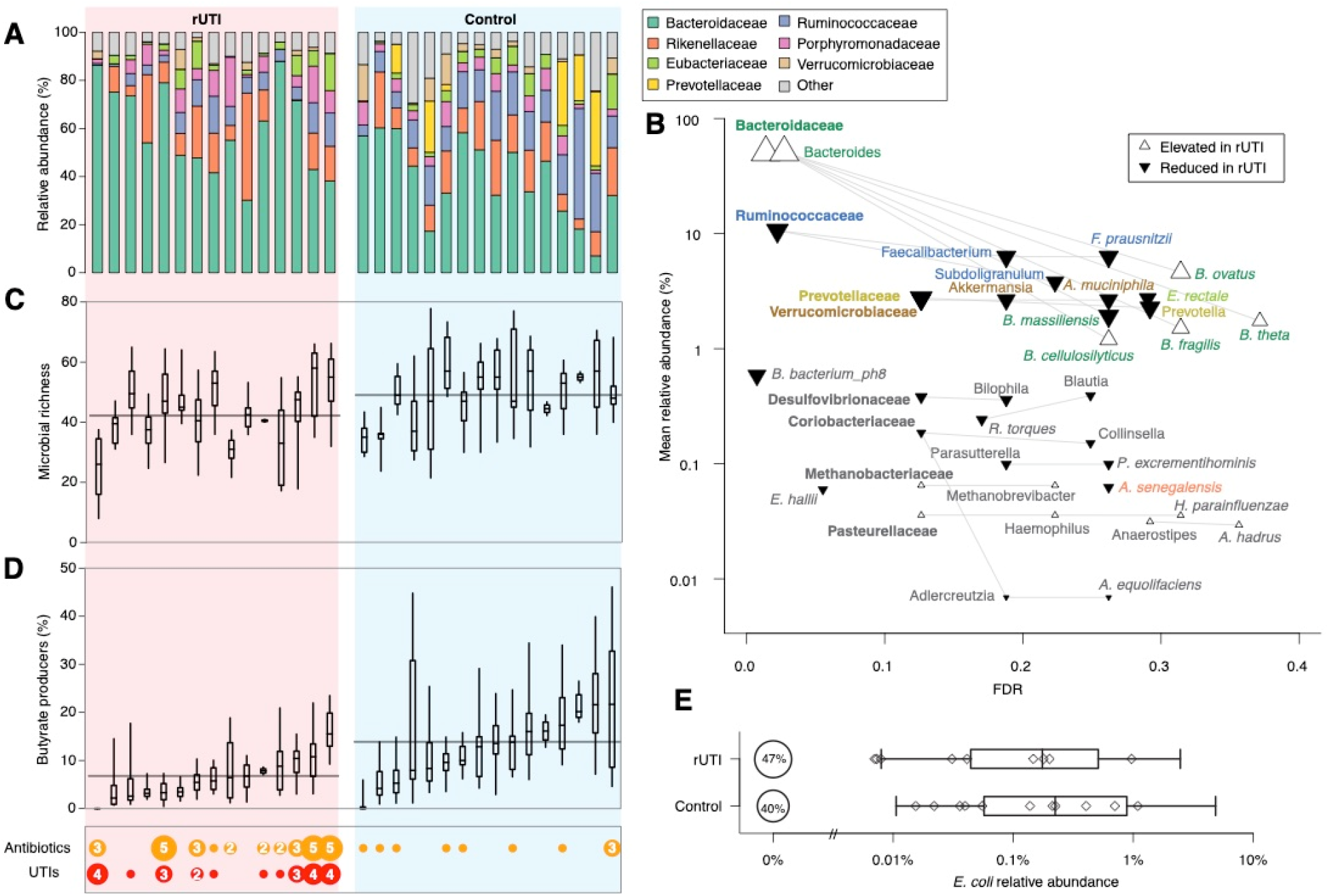
rUTI women have a distinct gut microbiome. (a) Average relative abundances of bacterial families for each patient in the rUTI (left) and the control (right) cohorts. (b) Significance and rUTI group effect size for selected taxa. Each point represents one taxon; its effect size and direction (symbol) for rUTI vs. control, false discovery rate (FDR) and mean relative abundance across all samples. Taxonomic relationships are represented by lines. FDR values calculated independently at each taxonomic level. Bold, regular and italic text denote family, genus and species levels (c) Microbial richness distributions and (d) cumulative relative abundance of butyrate-producing species for each study participant. Box plots depict IQR and central 95% quantile for each individual. Horizontal lines represent group-level mean of individual means. Antibiotic use and UTI occurrence for each study participant is shown at the bottom left; symbol size and numerals denote the number of UTIs/reported antibiotic courses. (e) Relative abundance of *E. coli* in each cohort. Symbols denote median relative abundances of individual patients; box plots denote the interquartile range and 95% central quantile of non-zero values.

Several of the taxa reduced in the rUTI gut, including *Faecalibacterium, Akkermansia, Blautia* and *Eubacterium hallii*, are associated with short chain fatty acid (SCFA) production, including propionate and butyrate, which exert an anti-inflammatory effect in the gut through promotion of the intestinal barrier function and immunomodulation ^20,21^. *Blautia* was additionally identified as the only taxon significantly depleted at the time of UTI relative to non-UTI samples (FDR=0.01). Cumulatively, SCFA producers, particularly butyrate producers, were significantly less abundant in rUTI women (p=0.001) (Figure 2d; Figure S2). Four KEGG Orthogroups ^22^ representing components of butyrate production pathways were significantly reduced across the rUTI cohort (Table S4). Functional analysis with HUMAnN2 ^23^ additionally revealed distinctions between the cohorts. These included pathways associated with sugar degradation and biosynthesis of metabolite intermediates and amino acids, which were depleted in the rUTI cohort (Table S5), many of which were also found to be differentially abundant in a study of irritable bowel syndrome (IBS) patients with sugar malabsorption ^24^.

The loss of gut microbial richness, diversity, and butyrate-producing bacteria that we observed is also a hallmark of exposure to broad spectrum antibiotics, including ciprofloxacin ^25–27^, which was used to treat more than a third of UTIs in our study. Thus, we sought to determine whether antibiotic exposure similarly had an impact on the gut microbiome of women in this study, and whether antibiotic effects may contribute to the observed shifts in microbiome composition in rUTI women (‘rUTI dysbiosis’). Though antibiotic exposure in the previous two weeks was associated with a significant reduction in microbial richness (p=0.05), this antibiotic-mediated loss of richness was not sustained. Samples taken 2-6 weeks after antibiotic exposure were not significantly different from baseline levels (p=0.2). Furthermore, we saw no association between the reported number of antibiotic courses and average richness (Figure 2c), and no differences in the overall gut microbiome stability between cohorts despite more frequent antibiotic treatment among UTI women (Figure S3). While low numbers of antibiotic exposures prevented a robust comparison of the effect by antibiotic class, we observed no differences in richness or in the abundance of butyrate producers between rUTI women with different antibiotic exposures (Figure S4). While we did not detect a lasting impact from individual antibiotic courses, it is still possible that repeated antibiotic use over multiple years may have contributed to the observed rUTI dysbiosis.

### rUTI gut dysbiosis shares broad similarities with inflammatory bowel disorders and may indicate low-level gut inflammation

The depletion of butyrate-producing taxa and microbial richness, key characteristics of rUTI dysbiosis, are also observed in other gut inflammatory conditions including nosocomial diarrhea ^28^, IBS ^29^ and inflammatory bowel disease (IBD) ^20^, particularly Crohn’s disease (CD) ^30^, and thus may be indicative of gut inflammation in rUTI women. While IBD is a multifactorial disorder for which the causative role of gut microbes is still incompletely understood ^31^, mouse models have helped to demonstrate a causal relationship between gut dysbiosis and gut inflammation^32^. To further explore the overlap between the shifts observed in IBD and rUTI guts compared to controls, we compared our data to longitudinal gut microbiome data from adults with and without IBD as part of the Human Microbiome Project 2 (HMP2) study ^33^, which shared the same extraction and sequencing protocols as our study (*Methods*). Relative to each study’s control group, we found that the ten most significantly depleted species in the rUTI gut, including butyrate producers *F. prausnitzii* and *E. hallii*, were also depleted in the IBD gut. We further observed a significant overall correlation in the estimated change of species-level abundances associated with rUTI and IBD (Figure S5), suggesting more general similarities.

There were also some notable differences. *Bacteroides*, significantly elevated in the rUTI group, did not differ between cohorts in the HMP2 study (Figure S5), and were also decreased among IBD patients in other studies ^34^. *E. coli* was significantly elevated in IBD patients in the HMP2 study, but showed no difference in average relative abundance between our cohorts (Figure 2e). Indeed, diminished *Bacteroides* alongside elevated *Enterobacteriaceae* was also observed in patients with nosocomial diarrhea ^28^. Diarrhea, also a symptom of IBD, is associated with reduced gut transit time, which has been shown to enrich for organisms common in the upper gastrointestinal tract, including *Enterobacteriaceae* ^35^, at the expense of anaerobic organisms such as *Bacteroides* ^36^. As such, rUTI women with low-level inflammation and no diarrhea may lack the depletion of *Bacteroides* and elevation of *Enterobacteriaceae* observed in other inflammatory, diarrhea-associated conditions. It is also possible that the considerable differences in treatment regimens for these conditions; i.e. antibiotics vs. anti-inflammatories, contribute to divergences of a common underlying inflammatory signal.

### Differences in host immune response may dictate susceptibility to UTI

Beyond inflammatory bowel disorders, rUTI dysbiosis also shares similarities with immunological syndromes affecting distal sites. For example, depletion of butyrate producers has been associated with rheumatoid arthritis, a systemic autoimmune disease which can be partially ameliorated in animal models with oral butyrate supplementation ^37,38^. Patients with chronic kidney disease (CKD) also exhibit a similar dysbiosis to rUTI women, including reduced levels of *Parasutterella* and *Akkermansia*, the latter of which has been found to be inversely correlated with interleukin-10 levels, an anti-inflammatory cytokine ^39^. We hypothesized that rUTI dysbiosis may also have an immunomodulatory role, potentially eliciting a differential immune response to bacterial invasion of the bladder. Thus, we broadly explored immunological biomarkers from blood samples collected at enrollment and at UTI, quantifying (i) a Luminex panel of human cytokines, chemokines, and growth factors involved in inflammation and T cell activation, and (ii) cell types and the transcriptional activity of peripheral blood mononuclear cells (PBMCs) (*Methods*).

Of the 39 analytes included in the Luminex panel, one chemokine, plasma eotaxin-1, was observed to be higher in rUTI women vs. control women at enrollment. Plasma eotaxin-1 is associated with intestinal inflammation ^40^, and levels of eotaxin-1 are increased in colonic tissue of patients with active IBD versus healthy controls ^41^. Subsequent human eotaxin-1 ELISAs validated these results, highlighting an additional link to dysbiosis-driven perturbation of the immune state, though, since this result did not hold after adjusting for race, we could not rule out potential demographic confounders. Eotaxin-1 was also higher in blood plasma of rUTI women at the time of UTI vs. enrollment (p=0.04; Figure S6a).

Although our relatively small cohort size provided limited statistical power for such cross-sectional comparisons, PBMC RNA-Seq analyses revealed two differentially expressed genes (FDR < 0.1), *ZNF266* and the long non-coding RNA *LINC00944*, were upregulated in the PBMCs of the rUTI cohort compared to controls (Table S6). *ZNF266* has been previously linked to urological health, identified as a potential PBMC biomarker for overactive bladder and incontinence in women ^42^. *LINC00944* has been associated with inflammatory and immune-related signaling pathways as well as tumor invading T lymphocytes in breast cancer, and markers for programmed cell-death ^43^. A CIBERSORT analysis of cell types based on transcriptional profiles revealed that resting NK cells were significantly reduced at the time of UTI relative to baseline levels (p=0.02; Figure S6b). NK cells help suppress bladder infection by UPEC in mice ^44^, so the loss of NK cells in the periphery may suggest a migration to the bladder at time of rUTI.

### rUTI and control cohorts showed similar *E. coli* dynamics in both the gut and the bladder

Previous work has implicated gut dysbiosis and a depletion of butyrate-producing bacteria in enhanced susceptibility to gut colonization by pathogens, including *Salmonella* ^45^ and *C. difficile* ^46^. While we could not quantify *absolute* species abundances, we observed no significant difference in the average relative abundance of *E. coli* between cohorts in this study (Figure 2e) suggesting that the rUTI dysbiotic gut is no more hospitable to *E. coli* colonization than controls. We considered the possibility that a temporal increase, or bloom, in *E. coli* relative abundance is a rUTI risk factor. Of the samples collected in the 14 days preceding an *E. coli* UTI, 75% exhibited *E. coli* relative abundance at or above average levels in the gut (Figure S7). However, elevated *E. coli* levels were not predictive of UTIs; none of the 22 *E. coli* blooms (here, defined as *E. coli* relative abundance more than 10-fold higher than the intra-host mean) occurred in the two weeks prior to UTI diagnosis. Thänert et al. identified intestinal blooms of uropathogens preceding some UTIs, but similarly noted that blooms often occurred in the absence of infection ^7^, leading us to conclude that elevated levels of *E. coli* may facilitate transfer to the bladder, but such gut blooms rarely manifest in infection. However, without frequent urine collection, we cannot rule out asymptomatic bladder colonization on such occasions.

Though we did not detect differences in *E. coli* species dynamics, the *E. coli* species encompasses considerable phenotypic and phylogenetic diversity. *E. coli* strains are broadly delineated into evolutionarily divergent phylogroups ^47^ with UPEC strains most commonly arising from phylogroups B2 and D ^4,48^. We hypothesized that rUTI dysbiosis may manifest in a qualitatively different *E. coli* population in the gut, contributing to increased susceptibility to rUTI. In order to explore *E. coli* strain-level diversity within stool metagenomes, we applied StrainGE, a tool suite to characterize the strain(s) of a given, low-abundance species within complex microbial communities ^49^ (*Methods*). Patterns of strain carriage were broadly similar in the rUTI (Figure 3) and the control (Figure S8) cohorts. Both the number of strains per sample and the phylogroup distribution were comparable between cohorts (Figure 4, Figure S9). While the majority of identified *E. coli* strains (62%) were transient, observed in one sample only, 22% were ‘persistent’, observed in at least one quarter of their carrier’s samples. Persistent strains were more likely to originate from phylogroups B2 and D than others (p=0.01), regardless of cohort, and were even slightly more common in control women (OR=2.1 (0.9, 5.2), p=0.1), at odds with the hypothesis of differential colonization resistance to phylogroups associated with UPEC between cohorts.

**Figure 3.**
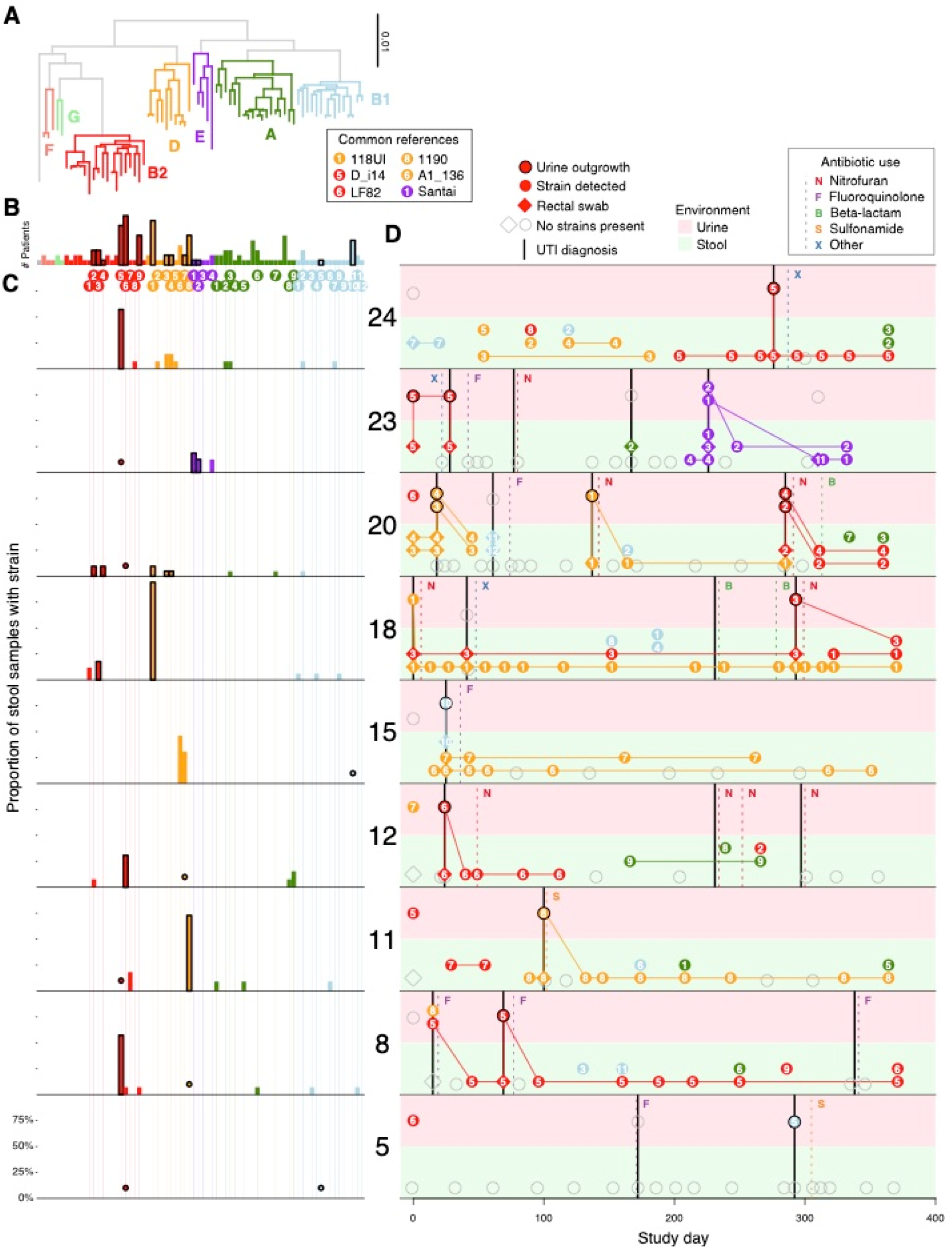
Frequent gut-bladder transmission and strain persistence in rUTI patients. Strain dynamics within all participants with *E. coli* UTIs. (a) Phylogenetic tree comprising strains called by StrainGE across all stool and urine samples, colored by phylogroup. (b) Number of unique patients with at least one strain observation, bolded if observed in any urine sample. Each strain identified in rUTI patients is uniquely identifiable by the phylogroup (color) and ID (numeral) indicated below. (c) Each panel represents one patient and the proportion of their stool samples in which each strain was identified; bold outlines if also observed in that patient’s urine. Dots represent observation in urine samples only. (d) Each panel represents longitudinal strain dynamics within one patient. Numerals refer to strain identifiers in (b). All strains are connected to their most recent previous observation in either fecal samples (green background), urine samples (pink). Diamonds denote clinical rectal swabs. Strains identified in urine outgrowth depicted if available (bold); otherwise raw urine strains are shown. Stool, urine or samples with no detected *E. coli* strains represented by open grey symbols. Vertical dashed lines represent self-reported antibiotic use, solid black lines denote UTI events.

**Figure 4.**
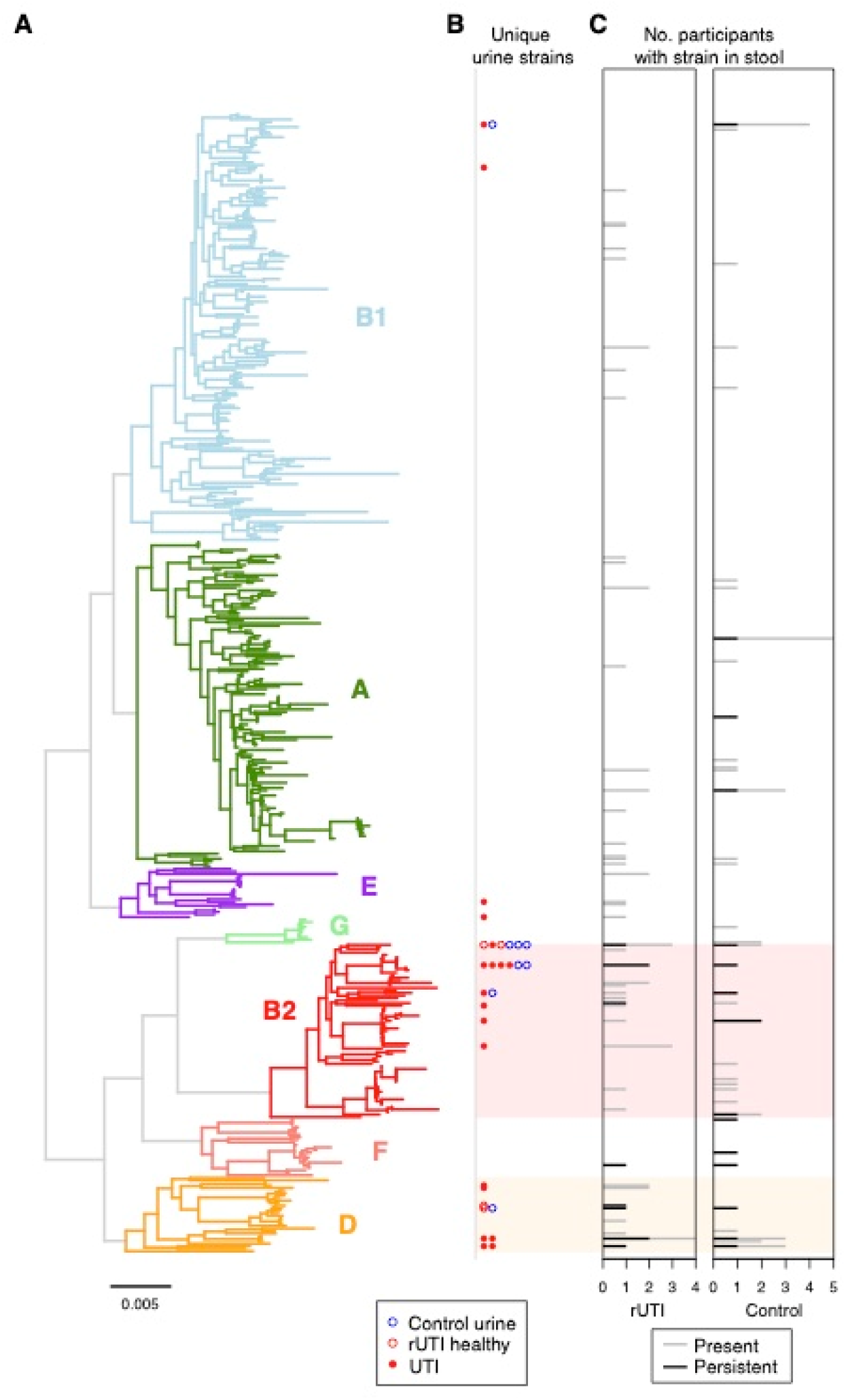
Phylogenetic distribution of *E. coli* strains identified in all stool and urine samples. (a) The phylogenetic tree of StrainGE reference strains colored and annotated by phylogroup. (b) Unique *E. coli* strains identified in urine samples are marked alongside the corresponding reference strain. Filled circles represent UTI-causing strains, blue circles denote strains identified in control hosts. (c) The total number of rUTI (left) and control (right) women with the corresponding strain present in stool samples. Black bars denote the number of women for whom the strain was persistent in the gut.

Given the similar *E. coli* strain dynamics observed in the gut between cohorts, we applied StrainGE to all urine samples, seeking to elucidate differences in strain dynamics in the bladder. We found that 79% (11/14) of *E. coli* UTIs were caused by phylogroup B2 (n=7) or D (n=4) strains (Table S2), approximately in line with previous studies ^4^. Of the 24 healthy enrollment urine samples yielding sufficient bacterial DNA to be sequenced and profiled (Table S7), we detected *E. coli* strains in 54% (13/24), including over half of samples (7/13) from control participants, despite the absence of symptoms. All but one of these strains also belonged to phylogroups B2 and D. Control urines carried *E. coli* strains that were phylogenetically similar to UTI-causing strains based on StrainGE predictions (Figure 4; *Methods*), despite divergent clinical outcomes.

Mapping urine metagenome assemblies to a curated virulence factor database showed that UTI-causing strains were enriched in virulence factors (including iron uptake systems (*sit, chu, iro, ybt* operons), colibactin (*clb*), and type 6 secretion systems) relative to an *E. coli* species-wide database, though many of these were also present in the one urine sample from a control participant for which we had sufficient coverage to assess gene content (*Methods*, Table S8). This apparent transition of a likely urovirulent strain to the bladder of healthy women without eliciting UTI symptoms is consistent with previous studies which have been unable to identify genetic markers of urovirulence in mice ^48^, or consistently discriminate between UTI and asymptomatic bacteriuria strains in women ^50^. Nevertheless, the divergence in clinical outcomes after bacterial invasion of the bladder may still arise due to phenotypic differences in *E. coli* strains reaching the bladder that are not readily apparent in genome comparisons. It is possible that rUTI dysbiosis could have an impact on UPEC gene expression; it has been shown that higher SCFA levels are associated with down-regulation of *E. coli* virulence factors including fimbrial and flagellar genes ^51^, however, such transcriptional analyses fall outside the scope of this study.

### rUTI strains colonize the gut and long-term clearance was not achieved by antibiotic treatment

While it is well known that UTIs are most commonly caused by UPEC resident in the gut, longitudinal dynamics of these strains within the gut are less well understood, despite the importance of such insights into developing rUTI prophylaxis. As such, we applied StrainGE to all urine samples to identify UTI-causing strains and their corresponding gut dynamics, in particular at the time of UTI, as well as after antibiotic exposure.

Four rUTI women suffered multiple confirmed *E. coli* UTIs, though only one was a same strain recurrence (individual 8) (Figure 3), suggesting that rUTIs in this study population were not frequently seeded by a monoclonal UPEC reservoir. Comparisons of sequence data from urine samples and cultured rectal swabs collected during UTI clinic visits revealed that nearly all (11/12) *E. coli* UTIs, for which we had corresponding same-day rectal swabs, contained the same UTI strain, underscoring frequent transmission between the gut and bladder. Though not a perfect proxy for native gut relative abundances, the dominant *E. coli* strain in 4 of the rectal swab outgrowths was not the UTI-causing strain, suggesting that some UTIs may be caused by minority strains. Only one UTI (individual 5, Figure 3) was caused by an *E. coli* strain never observed in any other sample from that individual. This phylogroup B1 UTI-causing strain likely arose from a source other than the gut, such as the urinary tract or the vagina, which have been implicated as UPEC reservoirs ^7,52^.

We anticipated that antibiotic exposure - particularly ciprofloxacin - would impact gut carriage of *E. coli* strains, and may explain the lower frequency of persistent colonizers in the rUTI group. Indeed, *E. coli* strains were detected by StrainGE significantly less frequently in stool samples provided in the two weeks following antibiotic use (OR=0.3 (0.13, 0.68); p=0.004). However, many strains apparently cleared by antibiotics were observed again at later time points; in fact, none of the UTI-causing strains observed in the gut was permanently cleared following antibiotic exposure. While low-level persistence that is undetectable based on the sequencing data is a possibility, we plated a subset of post-treatment stool samples onto MacConkey agar to culture *E. coli*. In many cases, we observed no growth, further suggesting the absence of these strains for a period of time post-treatment (Table S9). Furthermore, antibiotic susceptibility profiling of 12 UTI-causing strains isolated from proximate stool samples demonstrated that the majority were susceptible to the antibiotics to which they were exposed (Table S10). While a single stool sample is not completely representative of the gut microbiota, our evidence suggests that UTI-causing strains may be frequently reintroduced to the gut from alternative sources following antibiotic clearance of the bladder and gut.

### rUTI is linked to perturbations of the gut-bladder axis, highlighting potential for novel therapeutics

Our study design, data collection and culture-independent metagenomic sequencing approach allowed us to characterize dynamics of the gut-bladder axis in healthy and rUTI women. We propose that rUTI susceptibility is dependent, in part, on perturbation of the gut-bladder axis, which represents a previously undescribed syndrome, comprising gut dysbiosis and differential host immunology. We show that, compared to healthy controls, women suffering rUTI exhibited gut dysbiosis characterized by depleted levels of butyrate-producing bacteria and diminished microbial richness. This dysbiosis did not appear to impact *E. coli* dynamics within the gut; relative abundances and strain types were similar between cohorts, suggesting that gut carriage of urovirulent bacteria in itself is not a risk factor for rUTIs. Notably, *E. coli* was commonly identified in the urine of healthy women, including strains arising from UPEC-associated clades and harboring similar virulence factors. Based on our observations, rUTI gut dysbiosis is consistent with low-level gut inflammation, and is reminiscent of other disorders in which microbiome-mediated immunomodulation may play a role in disease severity. We identified several immunological differences despite the limited size of our cohorts, supporting the hypothesis of a differential host immunology. Our findings warrant further investigations to explore microbiome-host mucosal immune interactions in the context of rUTI susceptibility.

While identifying the origins of rUTI dysbiosis is outside the scope of this study, repeated antibiotic exposure is a highly plausible mechanism through which this dysbiosis is maintained. Whether the observed dysbiosis is the direct result of long-term antibiotic perturbation is unclear, and the relatively short study period precluded us from establishing such causal relationships here. In addition to the potentially detrimental impact of antibiotic use on the gut microbiota, we found that treatment also failed to clear UTI-causing strains from the gut in the long term. As such, mitigation of rUTI risk via manipulation of the *E. coli* population appears likely to require long-term therapy to overcome repeated introductions from non-gut reservoirs. rUTI treatment protocols targeting UPEC strains in the gut while resulting in minimal disruption to the remaining gut microbiota, such as small molecule therapeutics ^53^, may offer improved long-term prospects. While more evidence is required to fully characterize the causal mechanisms between dysbiosis and infection, our work highlights the ineffectiveness and potential detrimental impact of current antibiotic therapies, as well as the potential for microbiome therapeutics (e.g. fecal microbiota transplants ^10^) to limit infections via restoration of a healthy bacterial community in the gut.

## Methods

### Study design & sample collection

#### Enrollment

This study was conducted with the approval and under the supervision of the Institutional Review Board of Washington University School of Medicine in St. Louis, MO. Women from the St. Louis, MO area reporting three or more UTIs in the past 12 months were recruited into the rUTI study arm, while women with no history of UTI (at most one UTI ever) were recruited into the control arm via the Department of Urological Surgery at Barnes-Jewish Hospital in St. Louis, MO. We excluded women who: i) had inflammatory bowel disease (IBD) or urological developmental defects (e.g., ureteral reflux, kidney agenesis, etc.), ii) were pregnant, iii) take antibiotics as prophylaxis for rUTI, and iv) were younger than 18 years or older than 45 at the time of enrollment. All participants provided informed consent. A total of 16 control and 15 rUTI women were recruited to the study; 14 women in each cohort completed the entire study collection protocol. Participants who did not complete the study were included in cohort-level comparisons, but excluded from longitudinal analyses.

#### Sample collection & storage

Participants provided blood and urine samples, as well as rectal swabs, at the initial clinic visit. UTIs were diagnosed during clinic visits; additional UTIs (not presenting at the study clinic) were inferred based on symptoms and antibiotic consumption reported in the monthly questionnaire. Women visiting the clinic during the study with UTI symptoms provided rectal swabs, blood and urine samples, and were requested to submit stool samples as soon as possible (within 24 hours) after the clinic visit, as well as at a two week follow-up time point.

All participants provided monthly stool samples for 12 months. Samples were collected at home, and submitted via mail. Questionnaires were completed with all monthly and clinical sample collections; these captured self-reported antibiotic and drug use, dietary intake, sexual intercourse and UTI symptoms.

### Sample processing

#### Blood sample preparation

A total of 15 mL of blood was collected from each patient during initial enrollment and UTI visits. The blood was stored on ice for less than 30 minutes and then mixed with an equal amount PBS with 2% fetal bovine serum (FBS). Peripheral blood mononuclear cell (PBMCs) were then isolated using SepMate PBMC isolation tubes (Stemcell Technologies) with Ficoll-Paque PLUS density gradient medium (Cytiva). Serum was collected during the PBMC isolation process and stored at −80C until use. PBMCs were washed with PBS plus 2% FBS and pelleted via centrifugation at 10,000 x g at room temperature for 5 minutes. PBMC cell pellets were then flash frozen and stored at −80C until RNA extraction.

#### Rectal swab and urine preparation

Rectal swabs were collected in the clinic and stored on ice for less than 30 minutes. Rectal swabs were washed in 2 mL of PBS. 1 mL of PBS was centrifuged at 10,000 x g at room temperature for 2 minutes and the PBS supernatant was removed. The bacterial/fecal pellet was then flash frozen and stored at −80C until DNA extraction. The remaining 1 mL was then used to make serial dilutions and then plated on both Luria Broth (LB) and MacConkey agar and incubated overnight at 37C to quantify colony forming units (CFUs). After bacterial enumeration, bacteria from MacConkey and LB plates were scraped to collect bacterial outgrowths. Bacterial cells were washed with PBS, pelleted at 10,000 x g at room temperature for 2 minutes, flash frozen and then stored at −80C until DNA extraction.

Mid-stream urine samples were collected in sterile containers and stored on ice for less than 30 minutes. 10 mL of urine was centrifuged at 10,000 x g at room temperature for 5 minutes. The resulting pellet was washed in PBS, pelleted again, and then flash frozen and stored at −80C until DNA extraction. 1 mL of urine was used to make serial dilutions and then plated onto both LB and MacConkey and incubated overnight at 37C to enumerate CFUs. After outgrowth, the plates were scraped to collect bacterial colonies, which were then washed with PBS, pelleted at 10,000 x g at room temperature for 2 minutes, flash frozen and then stored at −80C until DNA extraction.

#### RNA Extraction - PBMCs

RNA was extracted from stored PBMCs using TRIzol Reagent (cat. no. 15596-026 and 15596-018; Life Technologies), according to the manufacturer’s protocol. Briefly, 0.75 mL of TRIzol was added per 0.25 mL of sample and cells were lysed by several rounds of pipetting. Samples were incubated for five minutes at room temperature. Chloroform was added to the samples at the recommended concentration and samples were incubated shaking for 15 seconds and set to rest for 2-3 minutes at room temperature. After incubation, samples were centrifuged at 12,000 x *g* for 15 minutes at 4C. The aqueous phase was collected for RNA isolation. RNA was precipitated using 100% isopropanol and incubated at room temperature for 10 minutes, followed by centrifugation at 12,000 x *g* for 10 minutes at 4C. The precipitated RNA was washed according to the protocol using 75% ethanol and resuspended in RNase-free water. Extracted RNA was stored at −80C until further use.

#### DNA Extraction – Rectal Swabs & Urine

DNA was extracted from rectal swabs and urine samples plated on MacConkey agar using the Wizard Genomic DNA Purification Kit (Promega), according to the manufacturer’s protocol. Briefly, samples were resuspended in 600 uL of Nuclei Lysis solution and incubated at 80C for five minutes, then cooled to room temperature. RNase solution was added to samples and incubated for 15 minutes at 37C, then cooled to room temperature. 200 uL of Protein Precipitation solution was added to the RNase-treated sample, vortexed for 20 seconds, and incubated on ice for 5 minutes. After incubation, samples were centrifuged for 3 minutes at 16,000 x *g* and the supernatant was transferred to a 1.5 mL microcentrifuge tube containing 600 uL of isopropanol. Samples were gently mixed and centrifuged for 2 minutes at 16,000 x *g*. The supernatant was removed and the DNA pellet was washed with 70% ethanol. Samples were centrifuged for 2 minutes at 16,000 x *g*, ethanol was aspirated and DNA pellet was air-dried for 15 minutes. The DNA pellet was rehydrated with DNA Rehydration solution and incubated at 65C for 1 hour. Extracted DNA was stored at 4C for short-term storage and at 80C for long-term storage until further use.

#### WMS sequencing & sequence data processing

Libraries were constructed from extracted DNA from stool, urine, rectal swabs, and plate scrapes using the NexteraXT kit (Illumina). Then, libraries were sequenced on a HiSeq 2500 (Illumina) in 101 bp paired-end read mode and/or a HiSeq X10 (Illumina) in 151 bp paired-end read mode. Sequence data was then demultiplexed. Samples that were sequenced multiple times on different runs were pooled together. Reads were processed with KneadData (v0.7.2, https://huttenhower.sph.harvard.edu/kneaddata/) to remove adapter sequence and trim low base qualities (with Trimmomatic), as well as to remove human-derived sequences (by aligning to human genome with bowtie2).

#### Luminex assays

Custom Luminex magnetic bead assay kit was obtained from R&D systems (product LXSAHM). Analytes from Human Inflammation and Human T Cell Response panels were chosen for the custom kit of 39 analytes: CXCL1/GROalpha, IL-1alpha, M-CSF/CSF1, LIF, LTalpha/TNF-b, MIF, APRIL, CCL11/Eotaxin, CCL4/MIP-1b, CXCL8/IL-8, IFN-g, IL-1b, IL-11, IL-13, IL-17A, IL-18, IL-21, IL-27, IL-31, IL-4, IL-6, MMP-1, TNF-a, BAFF/BLyS, CCL2/MCP-1, CX3CL1/Fractalkine, CXCL5/ENA-78, GM-CSF, IL-10, IL-12p70, IL-15, IL-17E/IL-25, IL-2, IL-22, IL-28A/INF-12, IL-33, IL-5, IL-7, MMP-3. Detection of the analytes in human plasma samples was performed using the Curiox DropArray system for miniaturization of magnetic bead multiplex kits. Plasma samples were diluted 2-fold for the assay. Results were read and quantified using a BioPlex multiplex plate reader and Microplate Manager software.

#### Eotaxin ELISA

Plasma eotaxin (CCL11) levels from rUTI and control patients were measured using the Eotaxin (CCL11) Human Simple Step ELISA kit (cat. no. ab185985; Abcam), according to the manufacturer’s protocol. Briefly, plasma samples were diluted into sample diluent and 50 uL of sample and 50 uL of antibody cocktail were added to 96 well plate strips. Plates were sealed and incubated shaking for one hour at room temperature. Wells were washed three times with 1x wash buffer and inverted to remove excess liquid. 100 uL of TMB substrate was added to each well; plates were covered to protect from light and incubated shaking for 10 minutes. Stop solution (100 uL) was added to each well and plates were incubated shaking for one minute. The OD_450_ was measured and recorded to determine the concentration of Eotaxin in pg/mL.

### Sequence data analysis

#### Community profiling and metrics

Bacterial community composition was determined using MetaPhlAn2 (v2.7.0 with db v20) ^54^ on KneadData-processed sequences. Functional profiling was performed by HUMAnN2 (v2.8.1, database downloaded in October 2016) ^23^ on KneadData-processed sequences. Diversity metrics and Bray-Curtis distances were derived from the MetaPhlAn2 relative abundance output using the *vegan* package in R [https://cran.r-project.org/web/packages/vegan/].

#### PBMC RNASeq analysis

Sequences from PBMC extracted mRNA were aligned to the human reference genome (hg19) using the STAR aligner ^55^. Picard-Tools (https://broadinstitute.github.io/picard/) was used to mark duplicate reads. Read counts per gene were generated with subread featureCounts ^56^. Read counts were normalized into Counts Per Million (CPM) using edgeR ^57^. This normalized read count matrix was then used as input for CIBERSORT using the LM22 signature gene set^58^. Results from CIBERSORT reported the relative abundance of 22 different immune cell types, including both PBMC and non-PBMC cell types, and it was used to remove three samples that were contaminated with 5% or greater of non-PBMC cell types. The CIBERSORT filtered set of samples was used to perform differential gene expression analysis using DESeq2 ^59^. Baseline healthy control samples were compared to baseline rUTI samples. Due to limited sample numbers and potential confounding, we included only samples collected from caucasian women in this analysis. Results driven by single outlying data points were not considered.

#### E. coli strain profiling

In order to track *E. coli* strain dynamics we used Strain Genome Explorer (StrainGE) ^49^. We applied the StrainGST module of StrainGE to identify representative *E. coli* strains in all stool, urine and rectal swabs, using an *E. coli* reference database generated from RefSeq complete genomes, as detailed in that paper. Strains mapping to the same representative reference genome in this database typically have an ANI of at least 99.9%. To provide further evidence that same-strain calls from sample pairs from the same host were indeed matches, we ran the StrainGR module of StrainGE, which calculates alignment-based similarity metrics. We used benchmarked thresholds to determine strain matches; strain pairs with a common callable genome >0.5%, Jaccard gap similarity >0.95 and average callable nucleotide identity >99.95% were deemed matches.

#### Determination of UTI-causing strains

Urine samples provided at the time of UTI diagnosis were plated on MacConkey agar. Sequence data was generated from DNA extracted from uncultured urine, and/or outgrowth on selective media. The cause of UTI was deemed to be the most abundant uropathogen, using outgrowth data where available, uncultured urine otherwise. Species were determined to be uropathogens based on UTI prevalence studies, e.g. ^1^.

#### Determination of virulence factors

Urine metagenomes for which *E. coli* represented the dominant species were assembled using SPAdes ^60^. To detect virulence factors in *E. coli* references (see StrainGST section above) and assembled genomes from study samples, we used the Virulence Factor Database (VFDB) for *E. coli* and the type 6 secretion system (T6SS) database (SecReT6) in genome-wide BLAST+ searches. Though VFDB contains T6SS genes, we removed them in favor of the T6SS-specific database for a T6SS-specific analytical pipeline. Other VFDB hits from blastn were filtered for ≥90% identity and ≥90% coverage. All *E. coli* genomes were separated by phylogroup for enrichment analysis, where Fisher’s Exact test was used to determine the significance of virulence factor enrichment in a certain phylogroup. T6SS hits were filtered for ≥90% identity and ≥90% coverage and the system was considered present where at least 12 different adjacent T6SS genes were present. Again, an enrichment analysis was performed using Fisher’s Exact test to determine the significance of T6SS presence in certain phylogroups.

### Statistical testing & models

#### rUTI risk factors

We used questionnaire responses to determine if any dietary or behavioral factors were associated with rUTI. We first compared the proportion of participants in each cohort who responded positively to binary variables (e.g. dairy consumption, alcohol etc. in the previous two weeks) in more than 50% or responses, and used a Fisher’s Exact test to determine significance. We next fit mixed effects logistic regression models to determine temporal risk factors for UTIs. Samples collected within 3 days of UTI diagnosis were classified as ‘time of UTI’; this binary variable was fit as a function of host (random effects term) and each dietary or behavioral response variable collected in the questionnaire. Variables with limited or no variance were excluded.

#### Identifying differences at the cohort level and time of UTI

We fit mixed effects linear regression models to compare the structure, diversity and function of the gut microbiome between cohorts. An arcsine square root transformation was applied to relative abundance values. Features (transformed relative abundances, diversity, microbial richness) were fit as a function of host (random effects term), cohort (categorical variable), and terms for antibiotic use and race (categorical variable) to adjust for potential confounding effects. To assess change in relative abundances at relevant timepoints, we also fit models including covariates for ‘pre-UTI’ (14 days preceding UTI diagnosis), ‘time of UTI’ (three days either side of UTI diagnosis), or post antibiotics (<14 days post antibiotic exposure) as binary variables. All taxa with more than 10% non-zero values were fitted using the *lme4* function in R. Wald’s test was used to derive raw p-values, which were adjusted for multiple hypothesis testing using Bonferroni-Hochberg correction at each taxonomic level.

The relative abundance of SCFA producers was additionally compared between cohorts; butyrate- and propionate-producing species were determined based on functional capacity to produce butyrate and propionate ^61^. These species’ relative abundances were then aggregated and compared as above.

We compared the stability of the microbiome between cohorts by assessing the distributions of within-host pairwise Bray-Curtis (BC) dissimilarities between individuals. Since rUTI women had, on average, slightly more frequent sampling than control women, due to the additional follow-up samples after UTI diagnoses, this metric may be biased towards smaller values in this cohort. However, we observed no significant trend between BC dissimilarity and time between samples, suggesting no detectable long-term trends. Furthermore, we detected no difference in the distribution of time-adjusted BC distances (BC divided by number of days between samples) between cohorts.

#### IBD comparisons

To compare rUTI dysbiosis to an IBD gut state, we downloaded MetaPhlAn2 output from the HMP2 study ^33^, (ibdmdb.org). We extracted longitudinal samples from adults with IBD (diagnosis=‘UC’ or ‘CD’) and non-IBD controls (diagnosis=‘nonIBD’). We fit linear mixed effects models with standardized relative abundances as a function of host (random effects term), race (race=‘white’; binary term) and recent antibiotic use. Fitted coefficients for the IBD and rUTI cohorts are then plotted in Figure S5.

## Data Availability

Sequence data are available from the Sequence Read Archive under Bioproject PRJNA400628.

## Acknowledgements

This project has been funded in part with Federal funds from the National Institute of Allergy and Infectious Diseases, National Institutes of Health, Department of Health and Human Services, under Grant Number U19AI110818 to the Broad Institute.

We would like to acknowledge members of the Broad’s Bacterial Genomics group and Hera Vlamakis for helpful conversations. We thank Brian Haas for assistance with PBMC RNA-Seq analysis as well as the Multi-Omics Core and Genomics Platform at the Broad Institute for sample processing and data generation.

## Author contributions

Study design HLS, KWD, SJH, AME

Study coordination HLS, KB, SBC, AK

Experiments performed HLS, JSP, CLPO, VLM, AEP

Data analysis CJW, HLS, TJS, LRvD, RAB, BO, BJH, CAD, WCC

Consultation and supervision of analyses BJW, ALM, TJH, TMH, ALK, HHL, KWD, SJH, AME

Prepared the original draft CJW, ALM, KWD, SJH, AME

Review and approval of the final manuscript was provided by all authors.

## Data availability

Sequence data are available from the Sequence Read Archive under Bioproject PRJNA400628.

---

**Figure S1.**
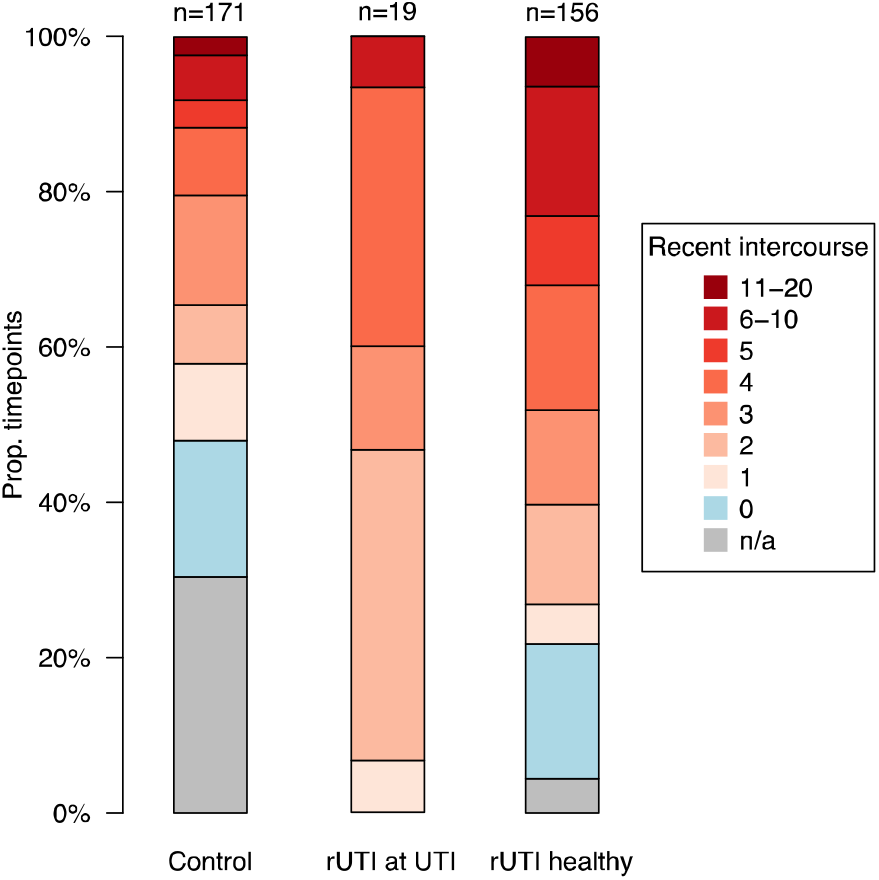
Sex precedes all clinical UTI events. Survey reports of intercourse frequency in the previous two weeks. Responses are partitioned by (i) control women, (ii) rUTI women at time of UTI, and (iii) rUTI women at non-UTI time points.

**Figure S2.**
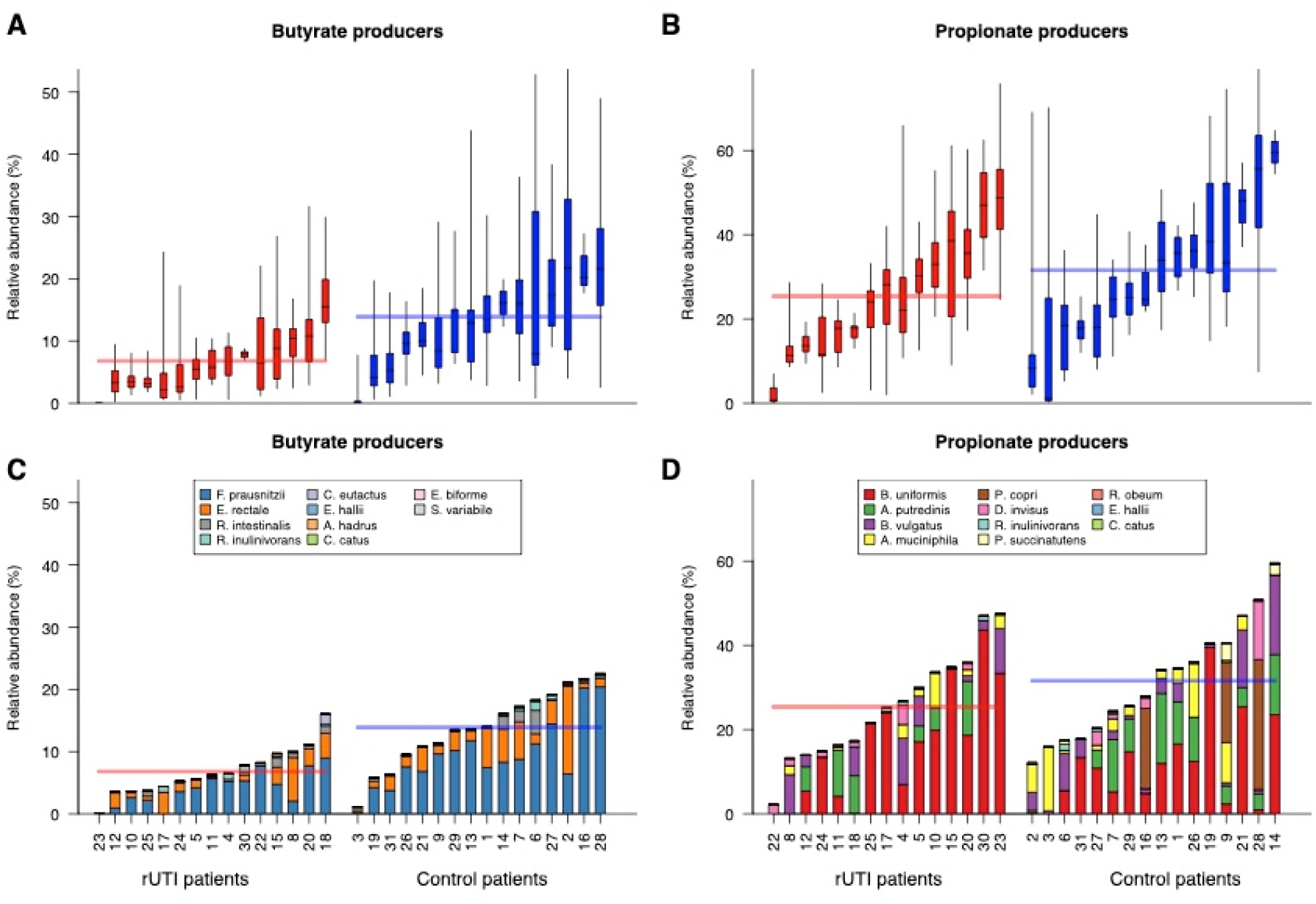
SCFA producing bacteria are depleted in the rUTI gut. Cumulative relative abundances of (a) butyrate and (b) propionate producing bacterial species in rUTI and control samples. Box plots denote the IQR and 95% central quantile. Within-host average relative abundances of individual species for (c) butyrate and (d) propionate producers are also shown.

**Figure S3.**
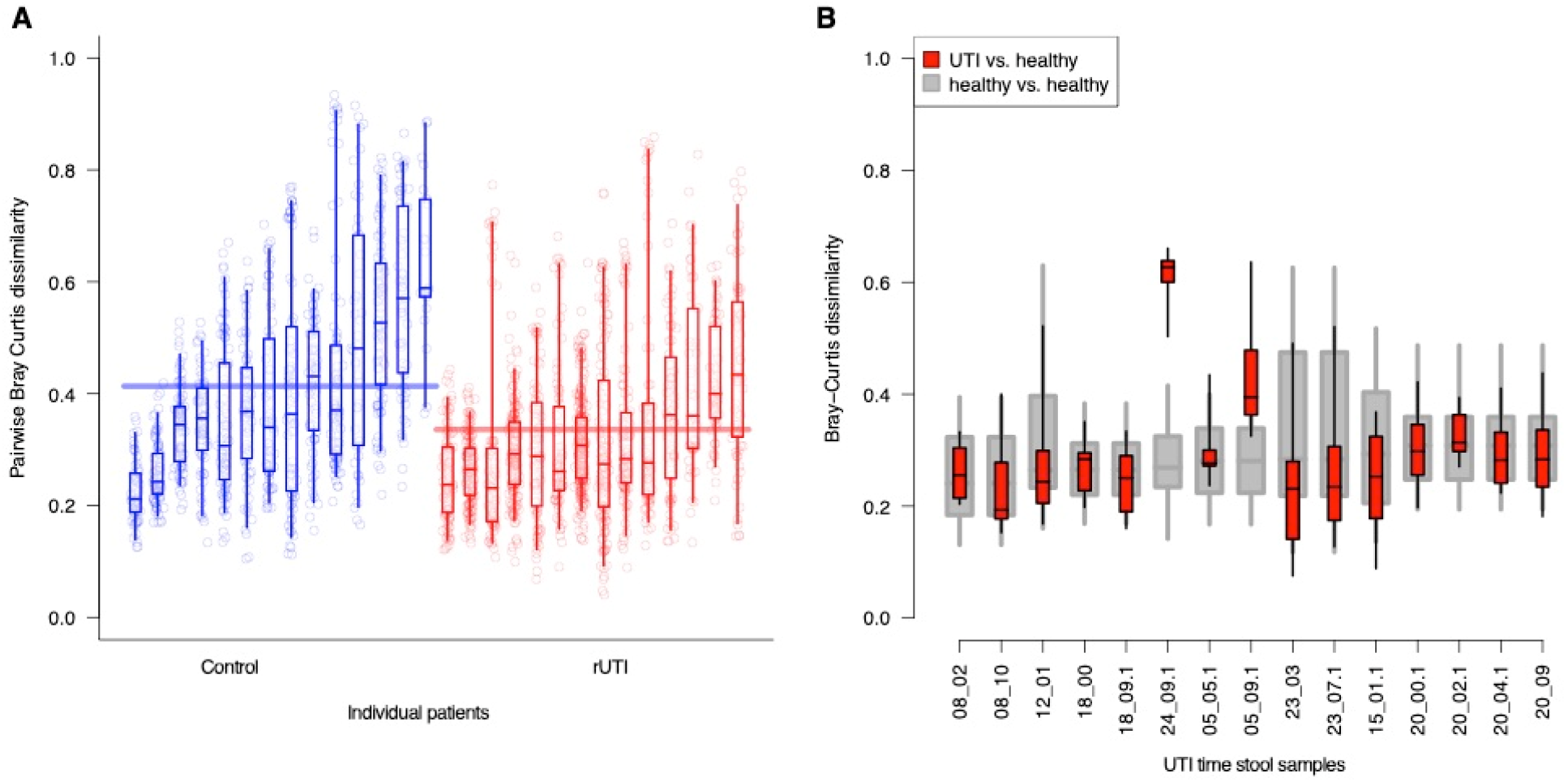
Bray Curtis dissimilarity across stool samples. (a) For each patient, the distribution of Bray-Curtis dissimilarities between all stool samples, ordered by increasing mean patient values within each cohort. (b) Bray-Curtis distributions between samples taken at the time of UTI vs. healthy time points (red), compared to all pairwise healthy sample comparisons. Box plots denote IQR & 95% central quantile

**Figure S4.**
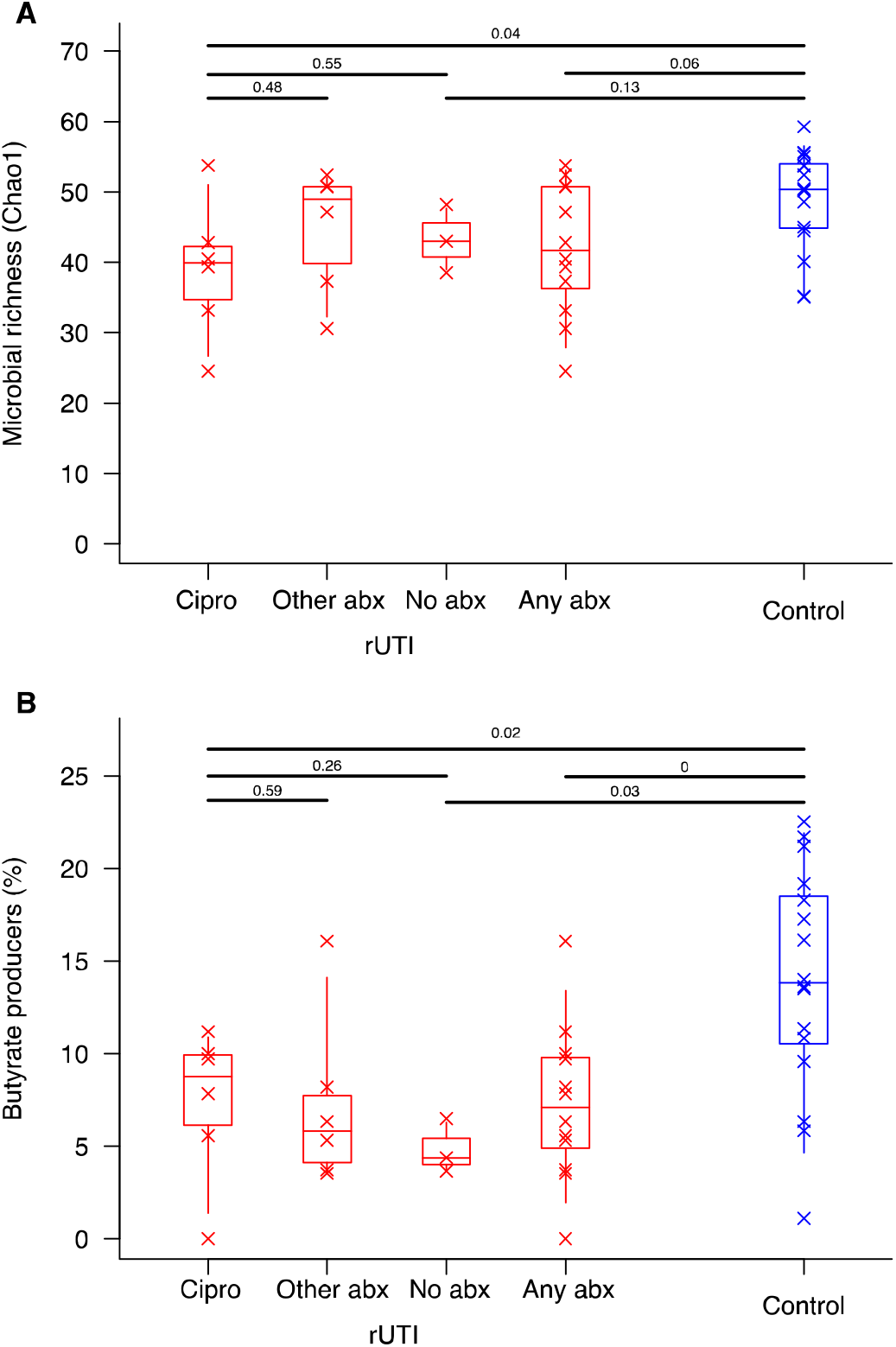
rUTI dysbiosis is not driven by antibiotic use during the study. We grouped rUTI women according to their antibiotic exposures at any point during the UMB study; (i) ciprofloxacin (ii) non-ciprofloxacin antibiotics; (iii) no antibiotics; (iv) any antibiotics. Groups were compared against each other and against the control cohort for (a) overall microbial richness and (b) relative abundance of butyrate producers. Crosses represent mean values for individuals, boxplots denote the IQR and 95% central quantiles for each group. Wilcoxon rank sum tests were applied to group pairs to derive p-values.

**Figure S5.**
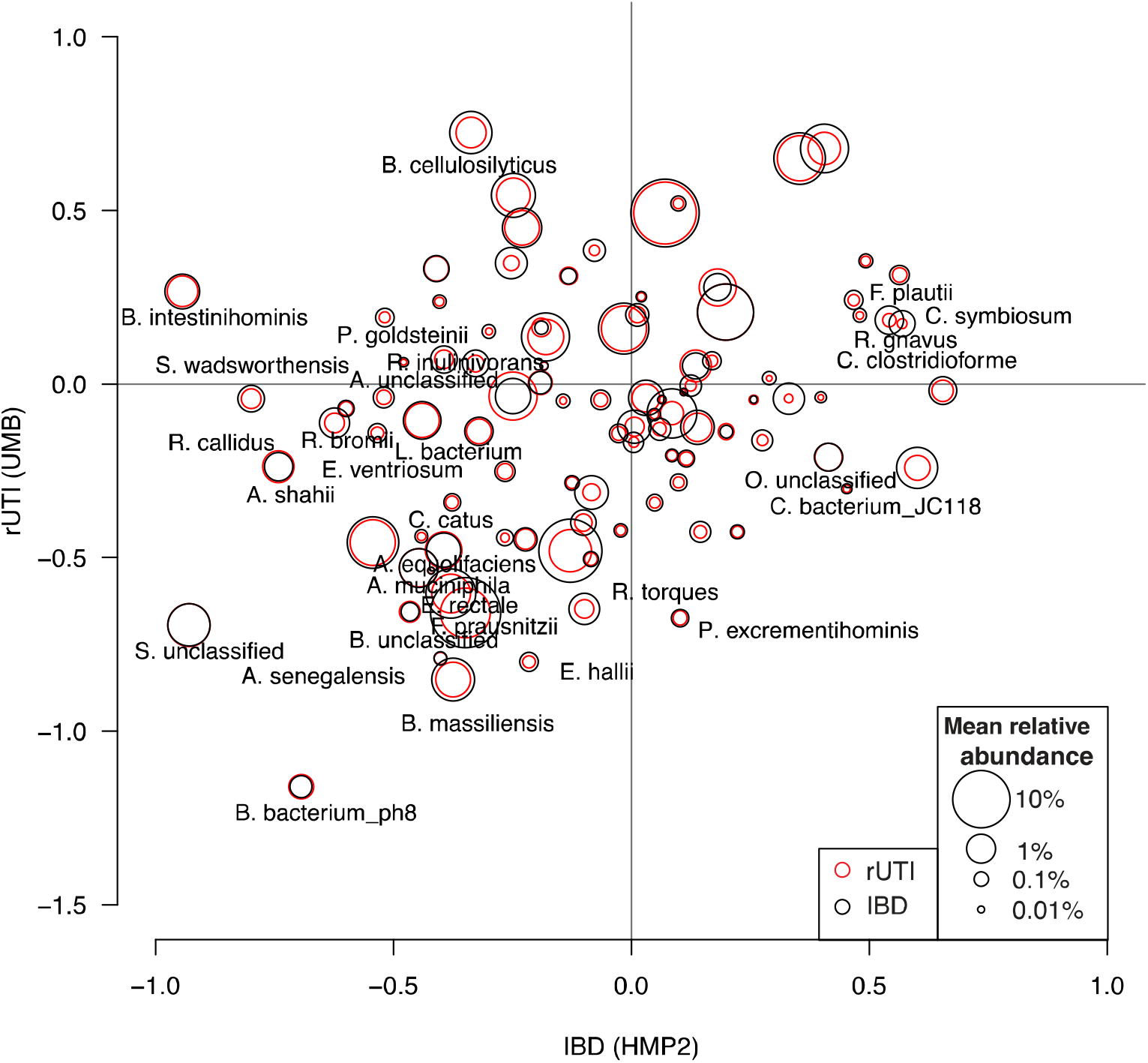
Most species depleted in the rUTI gut are also depleted in the IBD gut. We compared discriminatory taxa in rUTI women to those in IBD patients using data from adult participants in the HMP2 study ^33^. For each study, we fitted mixed effects models to standardized Metaphlan2 relative abundances as a function of categorical disease group (rUTI or IBD respectively, vs. each study’s control cohort), including covariates for race and antibiotic use. The disease group coefficients are plotted against each other for each species, with circle pairs representing the average relative abundance in each study. Species with uncorrected p values <0.05 in either study are labelled. Species not present in at least 10% of samples in either study are excluded. IBD comprises patients with either CD or UC.

**Figure S6.**
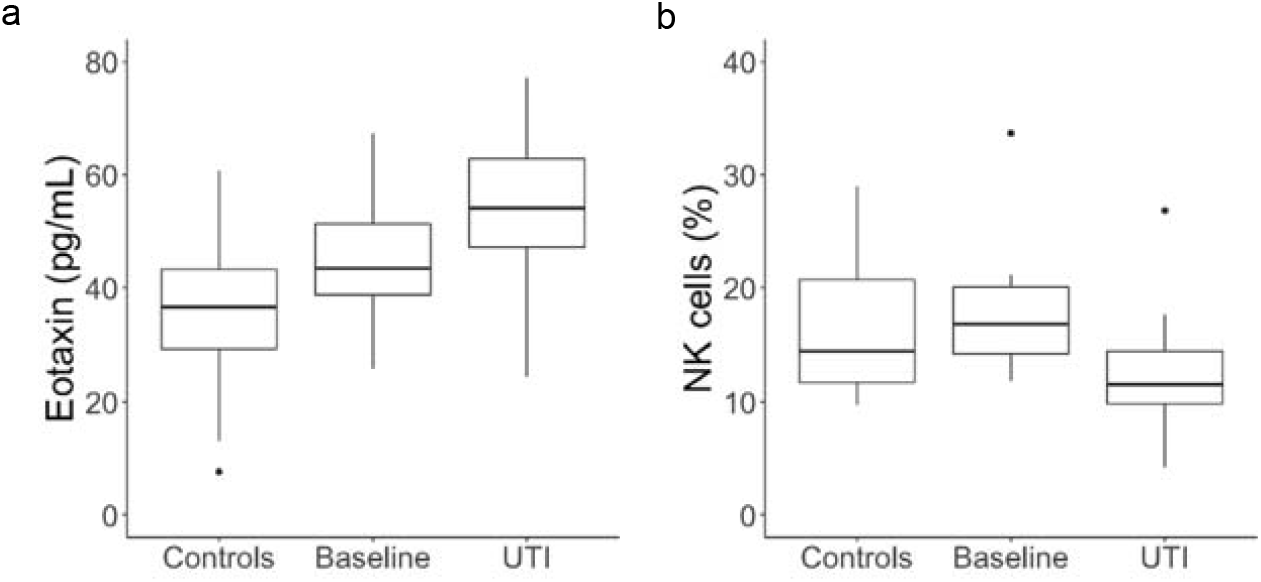
Immunological differences between cohorts. (a) Plasma eotaxin-1 levels in control women, and rUTI women at healthy enrollment and time of UTI. (b) Relative abundance of NK cells in control and rUTI women based on CIBERSORT output.

**Figure S7.**
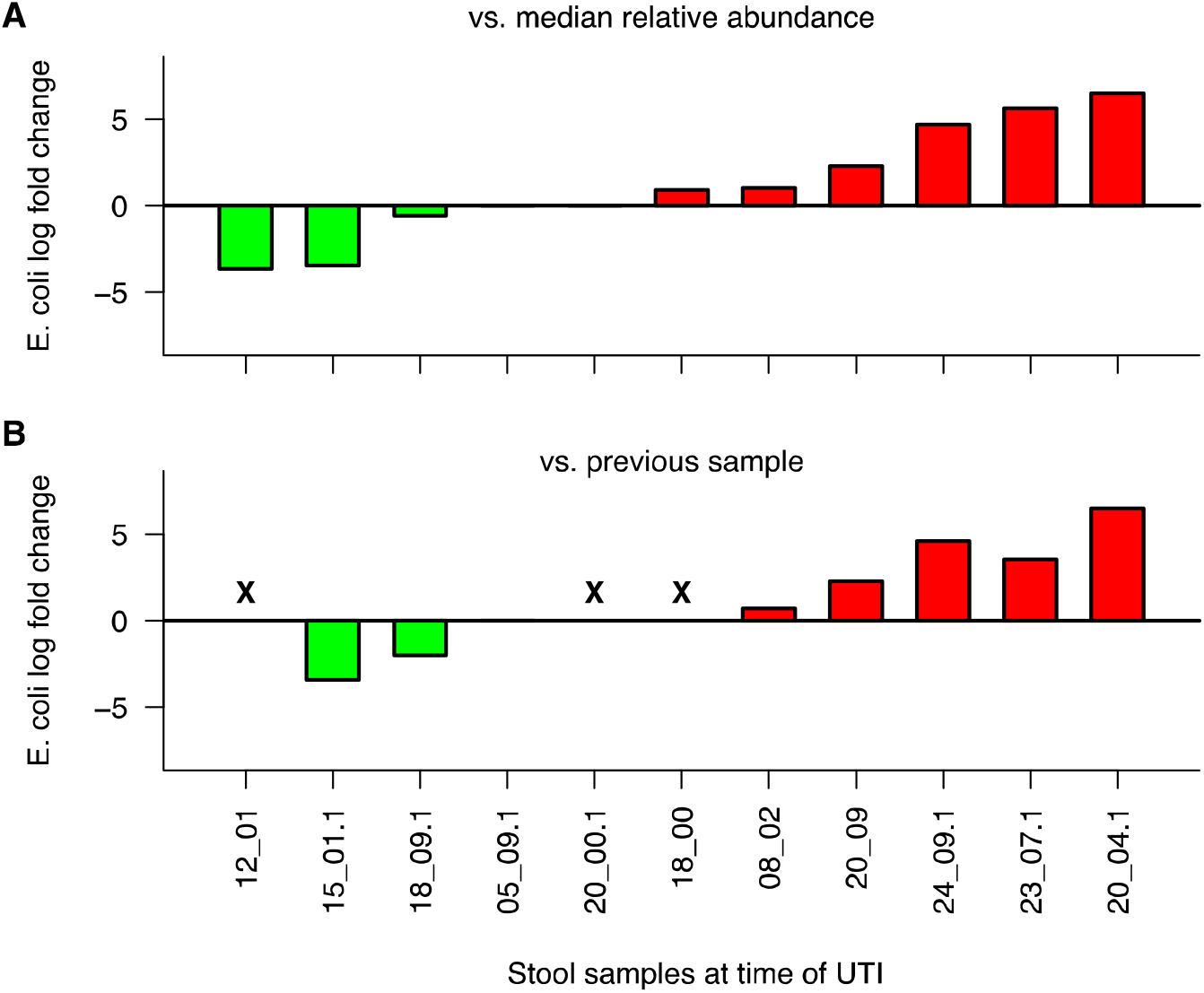
*E. coli* relative abundance around the time of UTI. For all stool samples taken within 3 days of a UTI event, the log fold change is given relative to (a) the median *E. coli* relative abundance in the corresponding patient, excluding samples taken at the time of UTI, and (b) the relative abundance of *E. coli* in the preceding stool sample. ‘X’ denotes samples for which there was no prior sample available.

**Figure S8.**
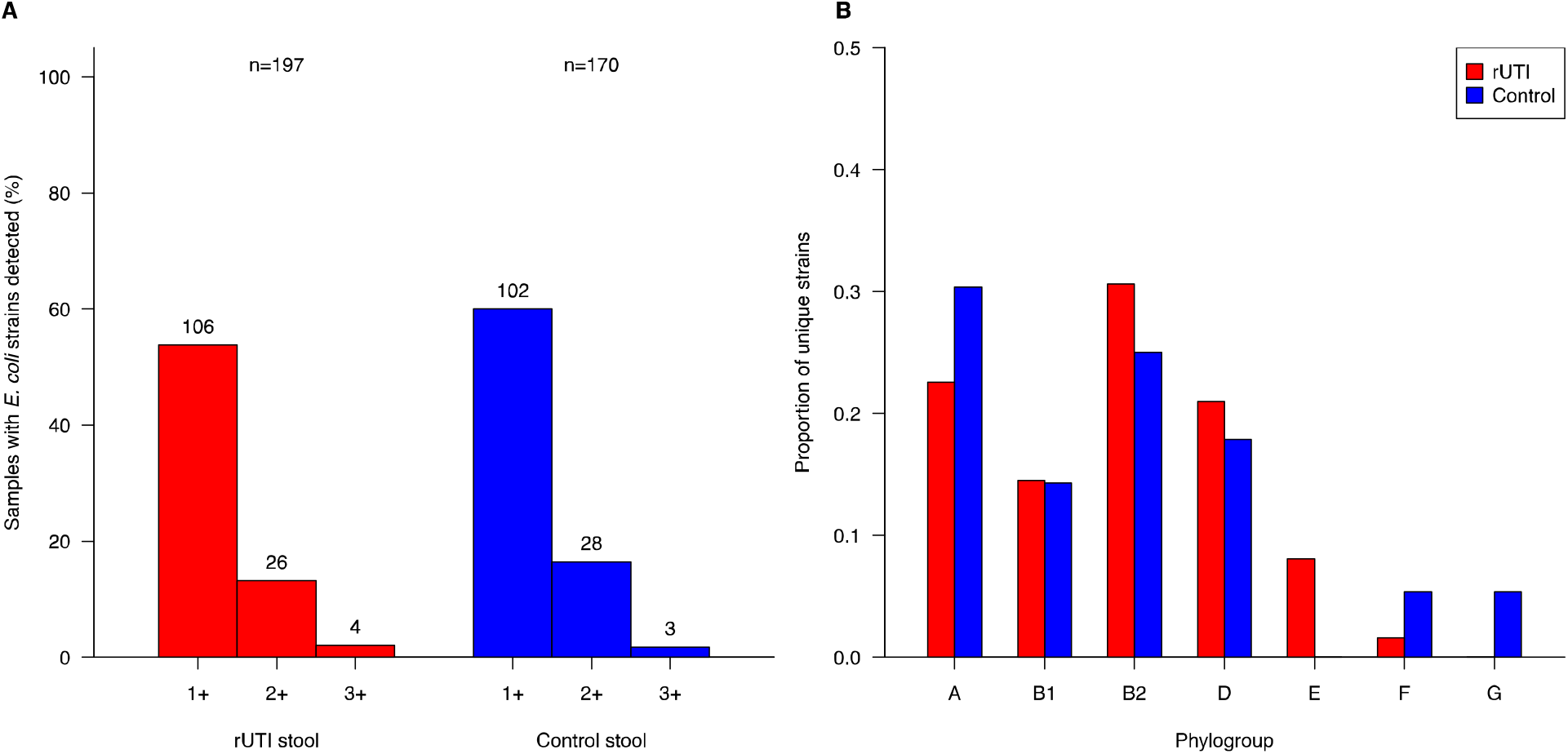
*E. coli* strain detection with StrainGE. (a) Number of detected *E. coli* strains by sample type. (b) Number of detected StrainGST reference strains vs. relative abundance of *E. coli*.

**Figure S9.**
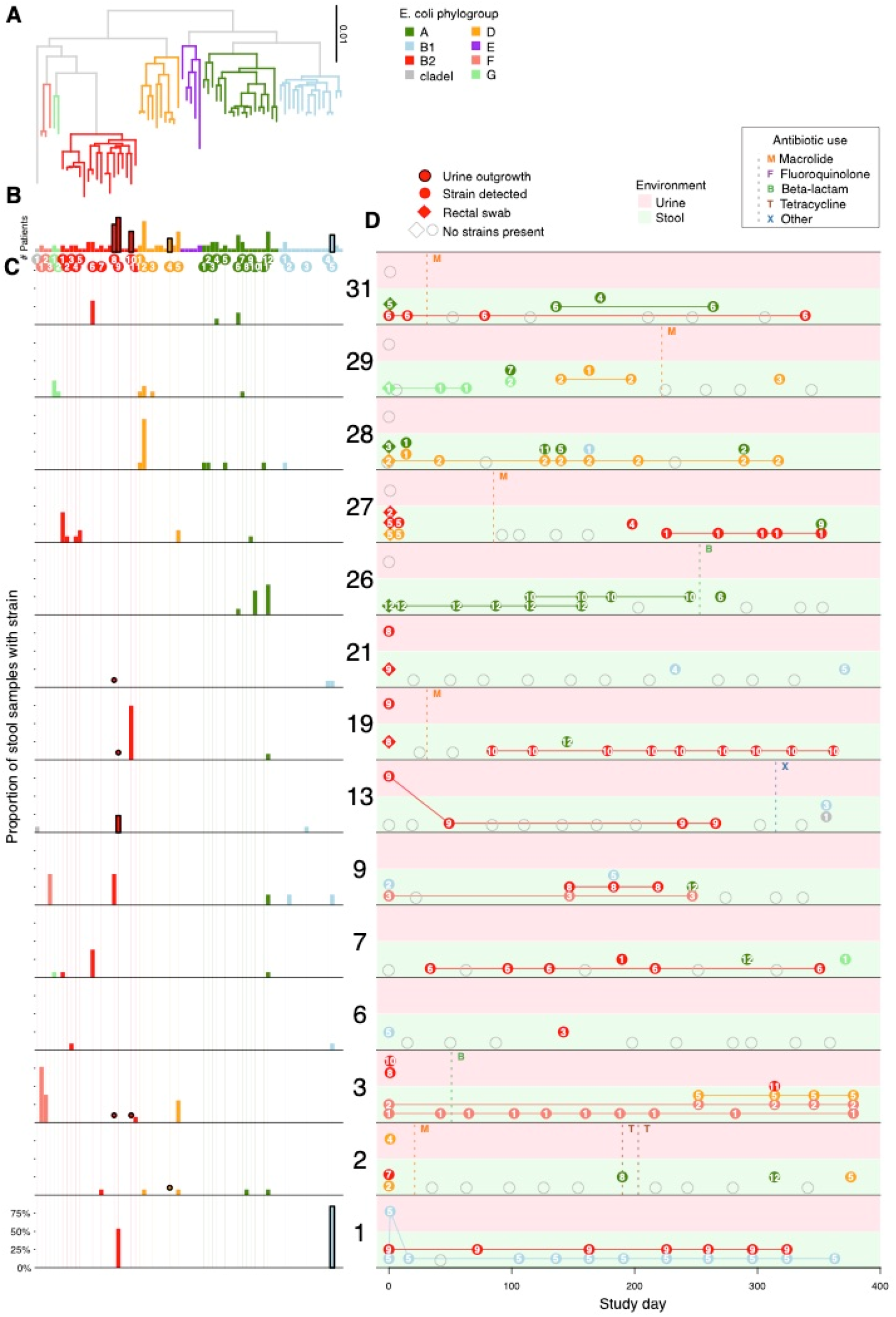
Strain dynamics in control women. Analogous to Figure 3 for the control cohort.

## Supplementary Tables

**Table S1. Cohort characteristics**. Demographic, behavioral and dietary characteristics of the rUTI and control women who completed the year-long study. Fisher’s exact test used to compare frequencies between cohorts.

**Table S2. Summary of UTIs and urine sequence data in the UMB study**. For each diagnosed UTIs, sequence data were available for cultured (plated on MacConkey agar) and/or uncultured urine samples. Metaphlan2 was used to estimate taxonomic composition and *E. coli* relative abundance. UTI species was determined to be the most abundant species associated with urovirulence. StrainGST provided representative *E. coli* reference strain(s), for which the corresponding clades and sequence types (STs) are also provided. The day of UTI is provided relative to the patient’s enrollment sample. Inferred UTIs did not have corresponding urine samples.

**Table S3. Differential taxa between rUTI and control cohorts**. At the species, genus and family levels, all bacterial taxa which differed between rUTI and control women with FDR<0.25. The fitted rUTI coefficient is estimated in a multivariable mixed effects regression model, adjusting for recent antibiotic use and race. Multiple testing correction applied separately at each taxonomic level.

**Table S4. Differential KEGG Orthogroups between rUTI and control cohorts**. Abundances of KEGG Orthogroups, derived with HUMAnN2, were compared across cohorts using linear mixed models. The fitted rUTI coefficient is estimated in a multivariable mixed effects regression model, adjusting for recent antibiotic use and race. All hits with FDR<0.05 are provided. KEGG Orthogroups belonging to butyrate production pathways ^62^ are marked.

**Table S5. Differential pathways between rUTI and control cohorts**. Abundances of pathways derived with HUMAnN2, were compared across cohorts using linear mixed models. The fitted rUTI coefficient is estimated in a multivariable mixed effects regression model, adjusting for recent antibiotic use and race.

**Table S6. Differential expression in PBMC RNA-Seq data**. Using DESeq, we sought differentially expressed genes between the rUTI and control cohorts. All genes with FDR<0.5 are listed.

**Table S7. *E. coli* in enrollment urine samples**. All urine samples collected at enrollment in the absence of UTI symptoms are described. The dominant species was determined by MetaPhlan2, and the relative abundance of *E. coli* strains provided by StrainGE.

**Table S8. Virulence factor enrichment in urine samples and *E. coli* database**. Virulence factors enriched in UTI urine samples vs. StrainGE *E. coli* database. Frequency across the 15 urine samples is provided alongside frequency in the database, partitioned by clades B2/D vs. non-B2/D. Fisher’s exact tests were performed to compare counts in urine vs. the *E. coli* database, as well as clade B2 vs. clade D within the database.

**Table S9. Outgrowths from stool samples post-antibiotic exposures**. A collection of stool samples for which *E. coli* strains appeared to have dropped out post antibiotic exposure were plated on selective media to determine if *E. coli* remained present at levels below detection limits for sequence data.

**Table S10. Antibiotic resistance across strains associated with bladder colonization**. *E. coli* strains were isolated from rectal swabs collected at enrollment and UTI diagnosis timepoints and submitted to antibiotic susceptibility profiling. Phenotypic resistance was compared to resistance gene content. Resistance breakpoints were: Nitrofurantoin ≥128ug/ml; Ciprofloxacin ≥0.12ug/ml; Amoxicillin ≥ 32ug/ml; Sulfamethoxazole-Trimethoprim ≥16ug/ml; azithromycin ≥32ug/ml (https://www.fda.gov/media/108180/download). Resistance determinants were derived using Resistance Gene Identifier ^63^.

